# Relationship between Body Mass Index, Gray Matter Volume and peripheral inflammation in patients with post-COVID condition

**DOI:** 10.1101/2024.10.25.24316095

**Authors:** Luise V. Claaß, Franziska Schick, Tonia Rocktäschel, Alejandra P. Garza, Christian Gaser, Philipp A. Reuken, Andreas Stallmach, Kathrin Finke, Sharmili Edwin Thanarajah, Martin Walter, Ildiko Rita Dunay, Bianca Besteher, Nils Opel

## Abstract

**Background:** Obesity is linked to low-grade peripheral inflammation and is recognized as an independent risk factor for severe COVID-19. Obesity and overweight have furthermore been shown to relate to structural brain alterations. Post-COVID condition (PCC) has in turn been associated with structural brain alterations and lingering immunological changes. Therefore, in this study, we aimed to assess whether obesity contributes to structural brain alterations and differences in immunological markers in PCC patients.

**Methods:** We investigated a previously established cohort of PCC patients (n = 61). Whole-brain comparison of gray matter volume (GMV) was conducted by voxel-based morphometry (VBM). Obesity, as measured by body mass index (BMI), as well as age, gender and total intracranial volume (TIV) were included as regressors in a linear regression model.

**Results:** A significant negative association was found between higher BMI and lower GMV in the right thalamus (p(FWE) = 0.039, k = 209, TFCE = 1037.97, x = 18, y = −21, z = 8). Moreover, BMI, GMV and immunological markers were linked in PCC. Specifically, BMI was primarily positively associated with Interleukin-6 and negatively with Interleukin-7, while GMV showed strong positive associations with Interleukin-8.

**Limitations:** A small cohort size and no available data on BMI changes before and after SARS-CoV2 infection limit the interpretation of our findings.

**Conclusion:** The results suggest that BMI contributes to GMV alterations in PCC patients, with both BMI and GMV being associated with peripheral immunological markers. These findings indicate that converging mechanisms, such as inflammation and structural brain alterations, may play a role in obesity and PCC.

## 1. Introduction

Elevated body mass index as a measure of obesity has extensively been linked to metabolic diseases and systemic low-grade inflammation [1]. Key mechanisms are likely the production of proinflammatory immunological markers and hormonal activity by adipocytes, perturbed glucose metabolism with insulin resistance, dyslipidemia, inflammation-associated endothelial dysfunction, and tissue hypoxia [1-7]. Furthermore there has been a broadened understanding of the significance and implications of obesity being one of the most prevalent somatic comorbidities of psychiatric diseases such as major depressive disorder (MDD) [8]. From this perspective, the way in which higher body mass indices are associated with structural brain alterations has been the subject of intensive research. In this context, patterns of cortical thickness reduction similar to those found in various neuropsychiatric disorders such as major depression and bipolar disorder have been observed in obesity [9].

Upon the beginning of the pandemic caused by severe acute respiratory syndrome coronavirus 2 (SARS-CoV2) obesity has been identified as a major risk factor for severe outcomes of coronavirus disease 2019 (COVID-19) [10, 11], possibly primarily conveyed by systemic chronic inflammation, dysregulated metabolism, dysfunctional immune system, inflamed endothelium, impaired mesenchymal stromal cells and altered adipose tissue [12]. Elevated pro-inflammatory immunological markers such as Interleukin(IL)-1β, Interleukin-6 and Tumor Necrosis Factor-alpha (TNF-α) have been linked to obesity [1] as well as to post-COVID condition (PCC), likely produced by overactivated monocytes/macrophages [13]. Furthermore, PCC has been linked to structural brain alterations. For instance, a large-scale longitudinal study by Douaud et al. found a reduction in gray matter thickness in the orbitofrontal cortex and parahippocampal gyrus [14]. Additionally, changes in diffusion measures, which serve as markers of tissue damage, were observed in regions functionally connected to the primary olfactory cortex. Patients with SARS-CoV-2 infection also exhibited a greater reduction in overall brain size [14]. In contrast, other preceding studies found wide-spanning larger gray matter volumes, parallel to lingering immunological changes, most pronounced in PCC patients with cognitive deficits [15, 16].

In sum, preliminary evidence suggests that both obesity and long-term consequences of COVID-19 show converging effects on cortical alterations and peripheral low-grade inflammation.

To our best knowledge, there has been no comprehensive investigation of the body mass index as modulating factor on immunological markers and structural brain alterations in patients with post-acute COVID-19 sequelae yet. Since obesity is a preventable risk factor, it would be worthwhile to understand the exact impact of this condition on PCC progression. Finding an increase in peripheral inflammation and cortical thickness alterations would explain the mechanisms by which obesity leads to severe courses of PCC. Additionally, it is our goal to include exploratory analyses to gain insights specifically into patients with more severe functional consequences of brain changes in regions critical to neuro-cognitive functions. We thus aim to enhance understanding of the factors contributing to the well-known heterogeneity among patient subpopulations affected by PCC [16].

## 2. Methods

### 2.1. Participants

The study participants belong to a previously established clinical cohort [16]. The study included 61 patients with PCC, recruited between April 2021 and June 2022 from the post-COVID outpatient clinic of the Department of Internal Medicine IV (Infectiology) and the Department of Neurology at Jena University Hospital.

One outlier in the VBM analysis was excluded from further analysis. As in the previously published report about this cohort [16], to examine a subpopulation with cognitive deficits in more detail, we divided the patients into two subgroups according to the result of the neurocognitive screening with the Montreal Cognitive Assessment (MoCA) based on a pre-specified cut-off of below 26 (mild to moderate cognitive impairment) and 26 or higher (no cognitive impairment) [17, 18].

For the final cohort the demographic characteristics are described in Table 1.

**Table 1.** Demographic characteristic of PCC cohort. SD = standard deviation, BMI = body mass index, MoCA = Montreal Cognitive Assessment.

|  | **Mean (SD)** | **Range (min - max)** |
| --- | --- | --- |
| **Age (years)** | 45.05 (12.93) | 20 - 73 |
| **BMI (kg/m<sup>2</sup>)** | 26.13 (4.43) | 18,21 - 36,13 |
| **Education (years)** | 11.14 (1.03) | 9 - 12 |
| **Time since infection (months)** | 9.94 (4.08) | 1 - 24,5 |
|  | **%** |  |
| **Male** | 40 |  |
| **Female** | 60 |  |
|  | **n** |  |
| **MoCA < 26** | 25 |  |
| **MoCA >= 26** | 35 |  |
| **Total** | 60 |  |

Initially, in each participant the acute SARS-CoV-2 infection was detected using real-time reverse transcriptase polymerase chain reaction (RT-qPCR) at the time of acute infection. At a later stage, a medical history was taken by a licensed physician. This included the timing and severity of COVID-19 symptoms according to WHO guidelines.

There were no restrictions on inclusion criteria regarding specific symptoms or time since infection to cover the full spectrum of post-acute sequelae of COVID-19. The frequency distribution of time since infection in our cohort can be seen in Figure 1. Participants had no history of psychiatric conditions, as verified by careful screening via MINI interview by trained personnel. In addition, none of the participants had a history of severe neurological disorders, relevant untreated medical conditions, acute infections treated with antibiotics, substance abuse or psychiatric disorders in first-degree relatives. All participants met the inclusion criteria for MRI examination and provided written informed consent to participate in the study.

**Fig. 1.**
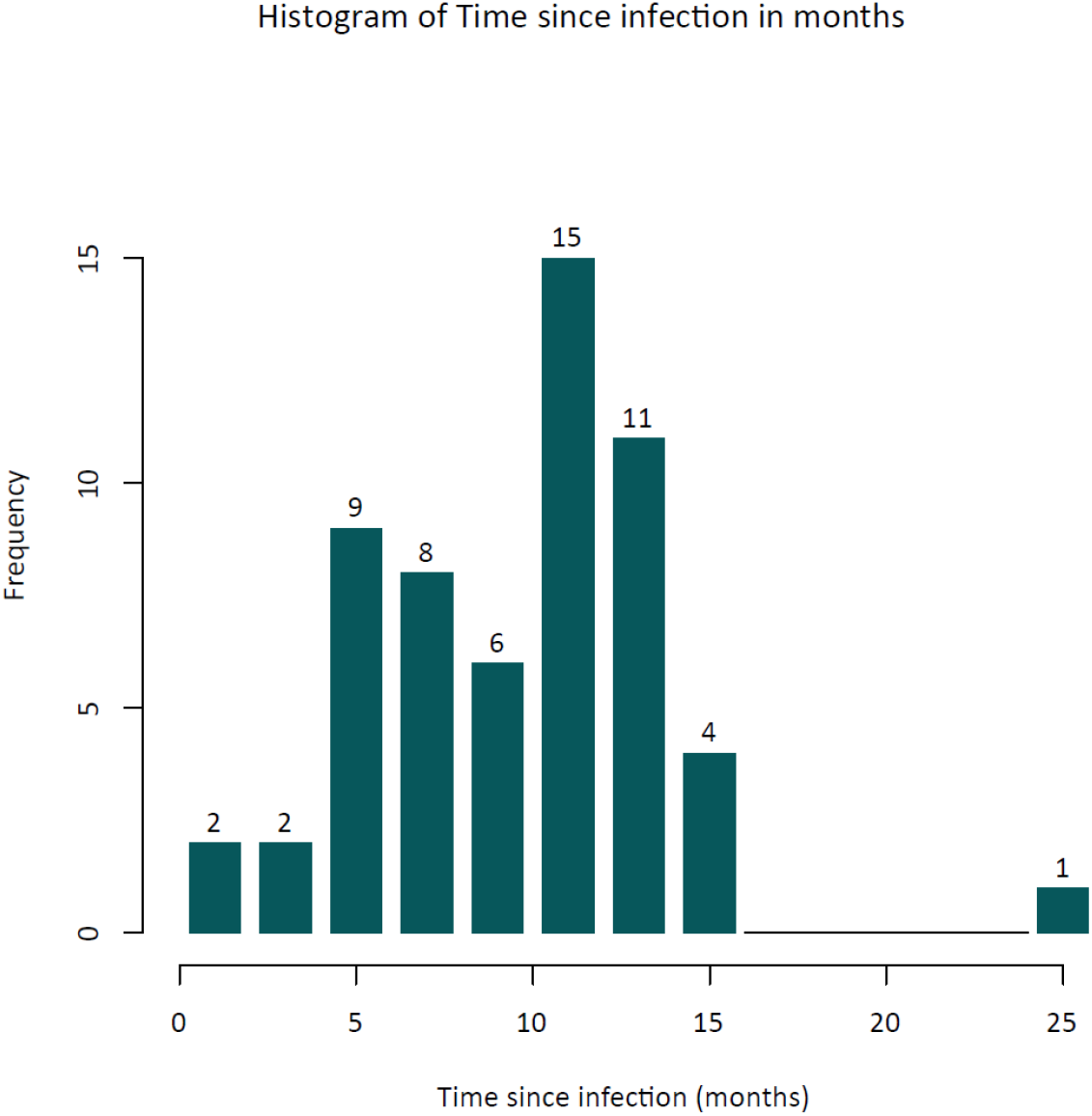
Frequency distribution of time since in infection in months. Numbers above the bars are absolute counts of patients.

The study protocol was approved by the local Ethics Committee of the Medical Faculty of the University of Jena.

### 2.2 Magnetic resonance imaging

Participants underwent a high-resolution T1-weighted magnetic resonance imaging (MRI) scan performed on a 3 Tesla Siemens Prism Fit scanner (Siemens, Erlangen, Germany). The scan was performed using a standard quadrature head coil and an axial 3-dimensional magnetization prepared rapid gradient echo (MP-RAGE) sequence with the following parameters: TR 2400ms, TE 2.22 ms, α 8°, 208 contiguous sagittal slices, FoV 256 mm, voxel resolution 0.8 × 0.8 × 0.8 mm. The acquisition time was 6 minutes and 38 seconds. Before the MRI scan of the native brain, all participants gave informed consent after a consultation with a licensed radiologist. This scan was part of a 60-minute MRI protocol. All scans were checked to ensure that no imaging artefacts were present.

### 3. Voxel-based morphometry

Voxel-based morphometry (VBM) was conducted using the CAT12 toolbox, developed by the Structural Brain Mapping group at Jena University Hospital in Jena, Germany [19]. This toolbox is integrated into the SPM12 software, which is produced by the Institute of Neurology in London, UK. All T1-weighted images were subjected to bias field correction. Subsequently, the images were segmented into gray matter (GM), white matter (WM), and cerebrospinal fluid (CSF) [20]. The images were spatially normalized using the DARTEL algorithm [21]. Furthermore, the segmentation process was extended to apply adaptive maximum a posteriori estimations [22] and to account for partial volume effects [23]. Following pre-processing, an automated quality check protocol was conducted to ensure the absence of artefacts. Subsequently, all scans underwent an 8 mm (Full Width at Half Maximum) Gaussian kernel smoothing procedure. To ensure the exclusivity of gray matter areas in the statistical analysis, an absolute gray matter threshold of 0.2 was applied. Data homogeneity and orthogonality were checked and outliers above a mean absolute z-score of 2 standard deviations were excluded.

### 4. Blood collection and serum levels of immunological markers

For the assessment of immunological markers, a standardized blood extraction was performed on all participants within the designated time frame of 7:30–9:00 a.m. on the day of the MRI scan. The participants were not fasted. 7.5 mL serum gel monovettes (Sarstedt) were used and 30 min after extraction, centrifugation was carried out at 1800 g for 10 min. The samples were divided into aliquots of 0.25 mL each and frozen at −80° C. Frozen samples were then transferred to our cooperating institute in Magdeburg, where serum was thawed at 4° C followed by a centrifugation step at 400 g for 10 min to remove any debris. The quantification of multiple key immunological markers including IL-10, IFN-γ, IL-6, TNF-α, CXCL10, sTREM2, sTREM-1, CCL2 (MCP-1), IL-18, BDNF, VEGF, β-NGF, sRAGE, CX3CL1, α-synuclein, G-CSF, IFNα2, IL-2, IL-7, IL-1RA and CXCL8 was performed using the LEGENDplex™ multiplex bead-based assay (BioLegend, 741091, 740795, 741197) following manufacturer’s instructions as previously described [24].

### 2.5 Statistical Analysis

#### 2.5.1. VBM

A multiple linear regression model, implemented in CAT12, was employed to study associations between BMI and whole brain gray matter volume in the cohort as whole. The model was performed on the whole-brain level, with BMI, age and gender included as covariates to remove related variance in all subsequent analyses. Total intracranial volume (TIV) as an index for head size was included as covariate as well, as there was no significant correlation between BMI and TIV in this patient group. An absolute threshold of 0.2 was used for masking. The results from the VBM were corrected using threshold-free cluster enhancement (TFCE) [25] with 5,000 permutations, and then corrected for multiple comparisons using the family-wise error method (FWE), with a significance level of *p* < 0.05. The anatomical labelling of clusters was conducted in accordance with the AAL atlas [26].

#### 2.5.2. Immunological markers

Statistical analysis and data visualization of serum immunological markers were conducted in R. Outliers were detected using z-score with a significance level of 0.05 and visual inspection in scatterplots. The data distribution was found to be non-normal, as indicated by Shapiro-Wilk tests. Consequently, Spearman’s rank correlation test was employed to assess the association between all immunological markers and both BMI and GMV values, to determine the presence of a linear relationship between BMI and specific immunological markers. To further evaluate the extent to which BMI or GMV could be explained by particular immunological variables, while adjusting for age and gender, multiple linear regression analyses were subsequently conducted. All statistical tests were executed with an alpha value of *p* < 0.05. Consequently, *p*-values of 0.05 or less were deemed statistically significant.

## 3. Results

### 3.1 VBM

The multiple linear regression revealed a significant correlation between BMI and a reduction in gray matter volume with a significant cluster in the right thalamus (p(FWE) = 0.039, k = 209, TFCE = 1037.97, x = 18, y = −21, z = 8), which is shown in Figure 2. Exploratory sensitivity analyses employing an uncorrected threshold of *p* < 0.001 and minimal cluster of k = 100 revealed several widespread clusters with GMV differences related to BMI. These clusters comprised regions including the left lobule IV of cerebellar hemisphere and right inferior parietal gyrus, excluding supramarginal and angular gyri (Figure 3).

**Figure 2.**
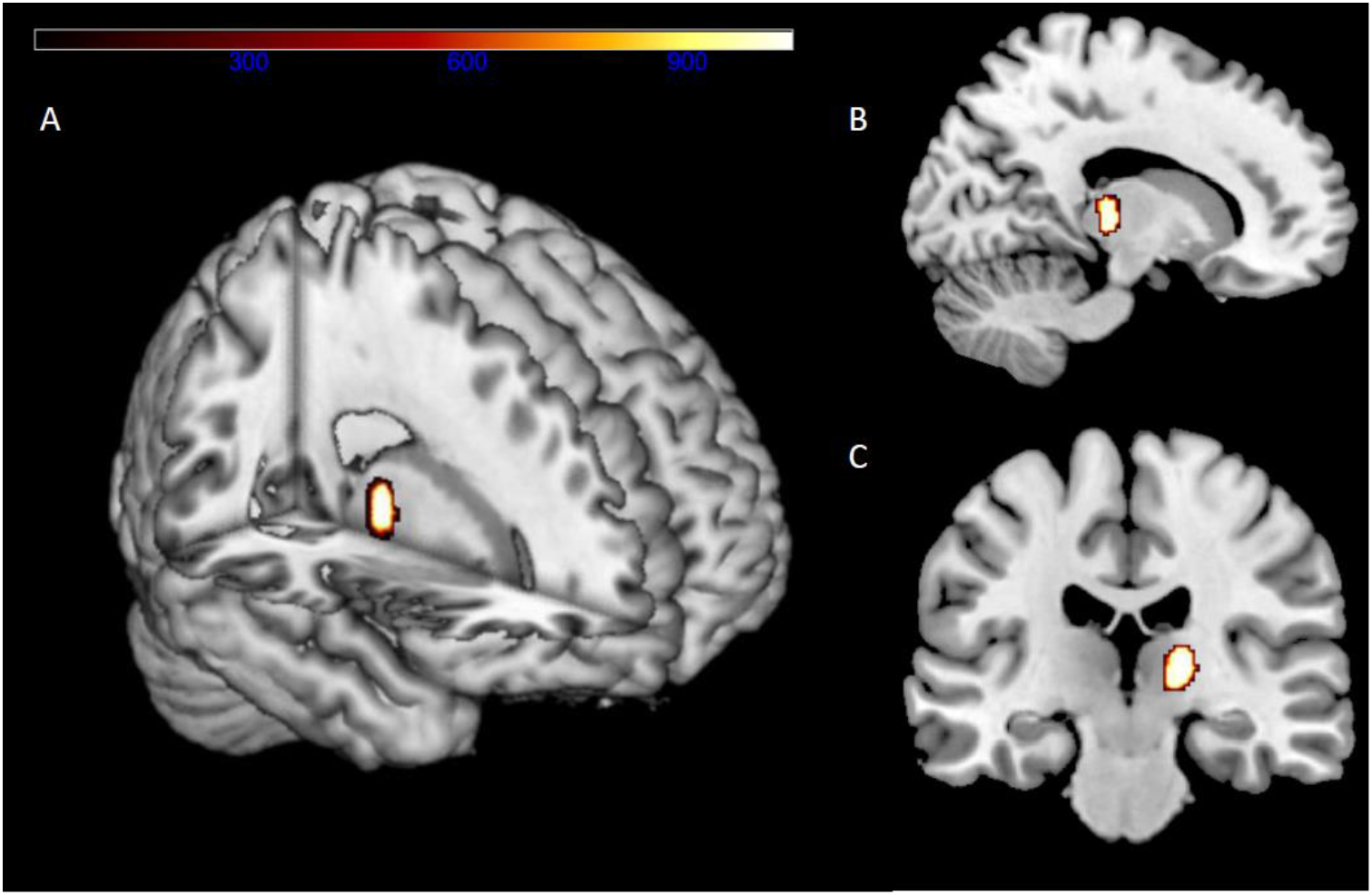
Reduced gray matter volume in patients with post-COVID condition correlates with BMI. Significant cluster in right thalamus (peak at MNI coordinates = 18-21 8, p(FWE) = 0.039) is presented as overlay. **A** 3D Render visualization of peak cluster. **B** Sagittal slice at x= 14, y= −27, z = 35. **C** Coronar slice at x = 34, y = −22, z = 26.

**Figure 3.**
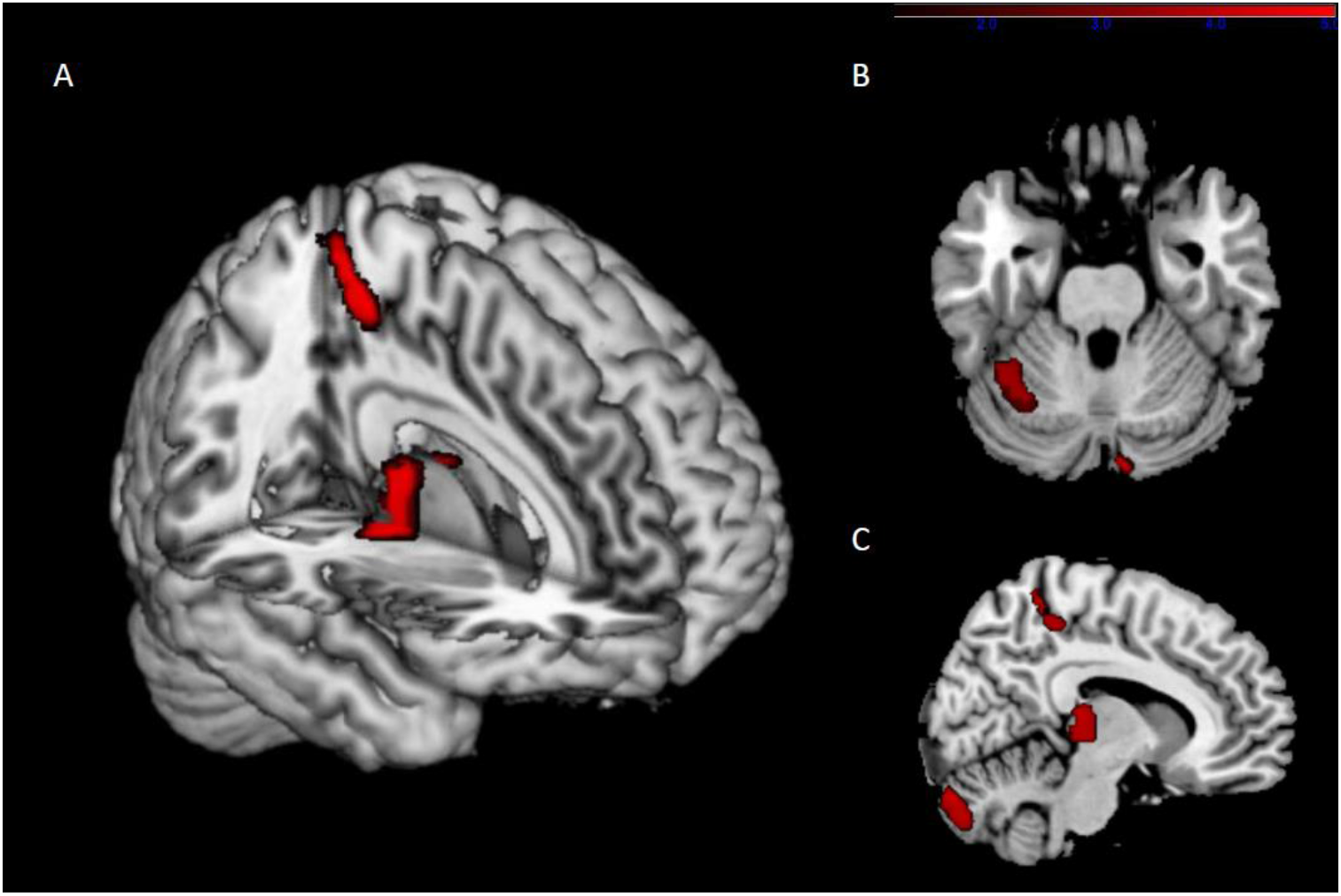
Exploratory sensitivity analyses employing an uncorrected threshold of *p* < 0.001 and minimal cluster of k = 100 showing several clusters with GMV reduction in relation to higher BMI. **A** 3D Render visualization. **B** Axial slice at MNI coordinates x = 3, y = 15, z = −26. **C** Sagittal slice at x = 9, y = 3, z = 32.

No positive association between BMI and gray matter volume was detected, neither at pFWE < .05, nor at the uncorrected exploratory threshold.

### 3.2 Immunological markers

Data on peripheral immunological markers were available for 50 of the 60 PCC patients included in the VBM analysis. Outlier detection led to exclusion of 2 subjects in the dataset.

A multiple linear regression model with BMI as dependent variable and immunological markers, age and gender as covariates yielded a significant effect (F(24,15)= 2.652, p = 0.027), indicating an positive association between BMI and IL-6 (p = 0,021) as well as a negative association with IL-7 (p = 0,021).

To further explore the relationship between levels of immunological markers and gray matter, we extracted gray matter volume information using the eigenvariate function in SPM12. The eigenvariate values of the cluster in the right thalamus as a spatially summarizing measure of gray matter volume were significantly positively correlated with IL-8.

A sensitivity analysis between IL-6, IL-7 and GMV revealed no significant association with time since infection, respectively. The coefficients for the correlations between immunological markers and BMI, as well as GMV can be seen in Figure 4A. Figure 4B illustrates the correlation matrix between these variables.

**Figure 4.**
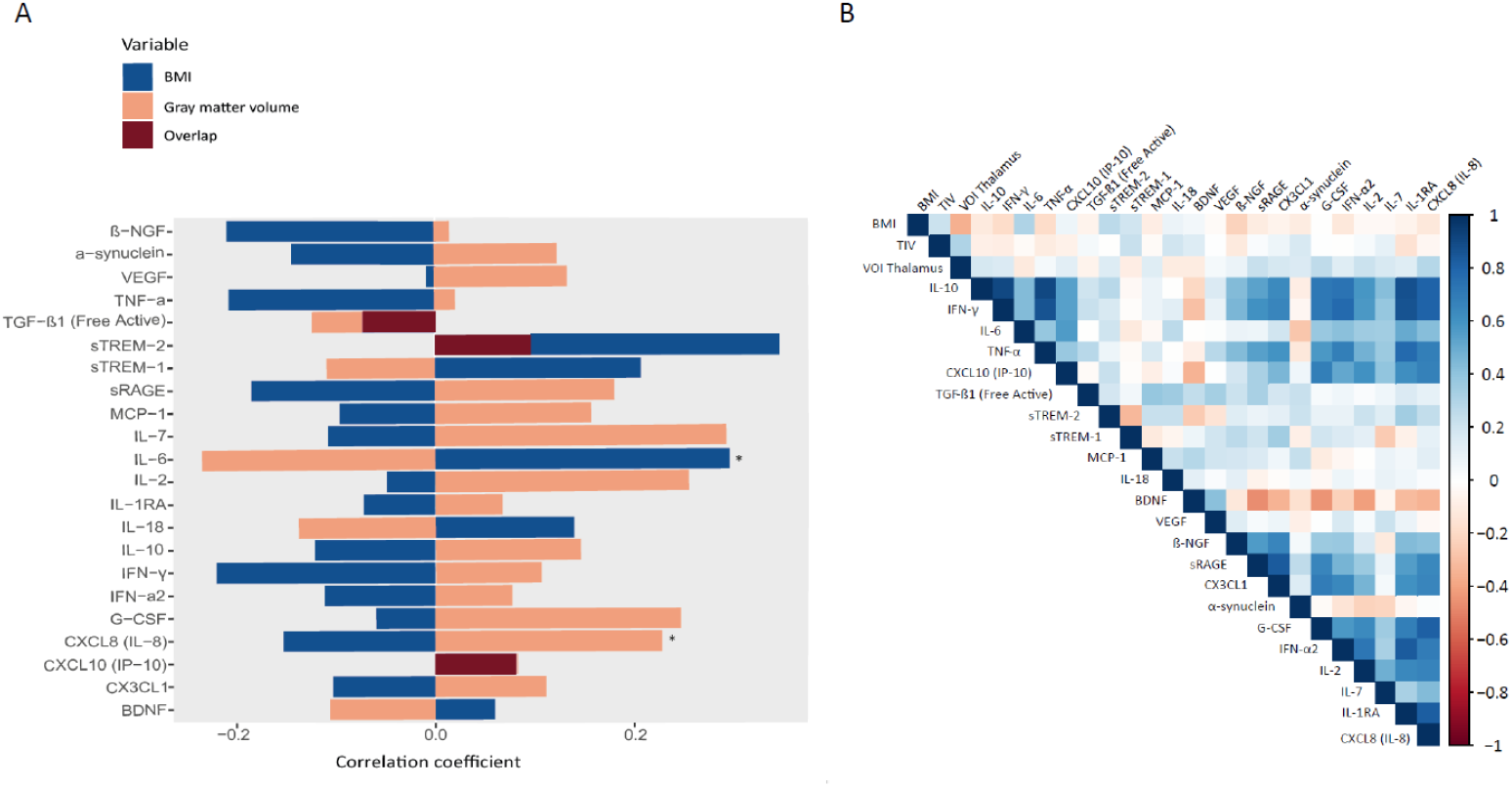
**A** Correlation coefficients between immunological markers and BMI, as well as between immunological markers and eigenvariate values of the cluster associated with gray matter volume reduction in patients with higher BMI. **B** Heat map of the correlation coefficients of BMI, total intracranial volume (TIV), eigenvariate values of the cluster in the right thalamus (VOI Thalamus) and immunological markers.

In the additional analyses to specifically examine a subgroup with cognitive deficits, we divided the patient cohort into 2 subgroups. The subgroup with reduced cognitive functioning comprised 25 patients (mean MoCA value 23.84, SD = 1.43) and the subgroup with normal cognitive functioning 35 patients (mean MoCA value 27.46, SD = 1.48).

The supplementary analyses exploring potential differences between these two subgroups revealed differing associations between immunological markers, BMI and GMV: While BMI was positively associated with IL-6 and negatively with Interferon-γ and TNF-α in participants with a lower MoCA score, BMI was predominantly positively associated with sTREM-1 in participants with a higher MoCA score. Regarding the eigenvariate values representing GMV, these were positively associated with Interferon-γ as well as Interleukin-2, G-CSF, Interleukin-8 and Interferon-α2 in participants with a lower MoCA score, while being correlated negatively with sTREM-1 in participants with a higher MoCA score.

## 4. Discussion

The goal of this study was to determine whether body mass index could impact the interactions between cortical volume and immunological markers in a cohort of patients with PCC. An association between BMI and gray matter volume was found, with a more pronounced GMV reduction being apparent in patients with higher BMI. These changes were most profound in the right thalamus, although an exploratory analysis suggested BMI-associated GMV changes were not restricted to a specific brain region. Furthermore, we found a significant positive association between BMI and IL-6, as well as a negative correlation with IL-7. The eigenvariate values of the cluster in the Thalamus were positively correlated with IL-8.

The existing literature documents widespread BMI-associated GMV changes in both healthy and neuropsychiatric populations [9]. To our knowledge, this is the first study to demonstrate these changes in a PCC patient group.

The thalamus, especially the paraventricular thalamic nucleus (PVT) has been linked to homeostasis of food intake and feeding behavior [27] and to the regulation of stress [28], so it is not improbable that its structure might be altered in obesity. In fact, previous volumetric imaging studies have found increased thalamic volumes in obesity [9, 29], whereas other studies using VBM found an inverse relationship between BMI or waist circumference and thalamic gray matter volume, respectively [30, 31]. This emphasizes the need to differentiate between measures of gray matter volume and density and underlines the difficulty of comparing measurements of gray matter in obesity obtained with different methods [32]. In the UK biobank study examining brain structure differences before and after an acute SARS-CoV2 infection, the strongest effect was observed in the volume of the thalamus when looking only at cross-sectional comparisons between the groups at the second post-infection time point [14]. However, when baseline scans were taken into account this effect disappeared, as the thalamus of participants who were later infected appeared to differ from that of healthy controls years before infection.

Regarding immunological markers, we found a significant positive association between body mass index and IL-6, which was still present after controlling for age, gender and time since infection. A multiple regression analysis also revealed a negative association between BMI and IL-7. In a previous study with the same clinical PCC cohort, no significant difference in the concentrations of these immunological markers was found between PCC patients and healthy controls [16]. When the cohort was divided based on the MoCA score as an indicator of cognitive functioning, BMI was significantly positively correlated with the proinflammatory immunological marker IL-6, and negatively correlated with the proinflammatory markers TNF-α and IFN-γ only in the group with the lower MoCA scores. Thus, rather than differentiating between all patients with PCC and healthy controls, IL-6 might have a relevant role particularly in patients with neuro-cognitive dysfunctions.

Of note, in a similar vein, results of Queiroz et al., who investigated cytokine profiles in acute COVID-19 infection and PCC, indicated that advanced age, comorbidities and elevated serum IL-6 levels are associated with severe acute COVID-19 cases and are good markers for distinguishing between severe and mild cases [33]. Moreover, IL-6 has been proposed as potential mediator of long-term neuropsychiatric symptoms of COVID-19 [34]. However, meta-analytic results suggested a relationship between the IL-6 pathway and weight regulation, with IL-6 pathway inhibitors being associated with increases in weight and BMI [35]. In a study linking cytokine profiles to symptoms of depression and anxiety, higher IL-6 levels were associated with decreased appetite. Additionally, the included Mendelian Randomization analyses indicated that genetically predicted higher IL-6 activity was associated with an increased risk of fatigue and sleep disturbances [36].

In our analysis correlating BMI-associated GMV changes with cytokine profiles, the cluster in the right thalamus was significantly positively correlated with Interleukin-8, which has been previously suggested as a biomarker to predict COVID-19 severity and prognosis [37]. After dividing the cohort based on the MoCA score, this correlation remained present only in the group with lower MoCA scores. Furthermore, GMV correlated with additional immunological markers, such as IL-2, which has been associated with a sustained immune humoral response and critical COVID-19 [38, 39].

Ultimately, it is known that obesity increases the risk of more severe disease progression in COVID-19; yet the underlying mechanisms remain unclear. A previous study demonstrated that SARS-CoV2 invades human adipose tissue, targeting both mature adipocytes and a subset of adipose tissue macrophages, which in turn leads to the activation of the immune system and the release of immunological markers associated with severe cases of COVID-19 [40].

Still, the practical implications of our findings are not yet fully decoded. In general populations, higher IL-6 levels seem to reduce appetite, whereas in COVID-19, elevated IL-6 levels are associated with greater disease severity. IL-6 inhibitors such as tocilizumab have shown protective effects in early stages of COVID-19 against the later onset of depressive symptoms [41], but they have also been linked to an increase in BMI when used to treat immune disorders [35].

Thus, anti-inflammatory treatment could not only benefit the disease course of severe COVID-19 and post-COVID conditions, but also have positive effects on obesity-related low-grade inflammation and its negative consequences. However, a potential worsening of obesity during treatment with cytokine inhibitors, such as tocilizumab, could also be expected.

A limitation of the study is the small cohort size and the lack of data on BMI changes before and after SARS-CoV2 infection, as this would allow us to account for possible weight loss or weight gain as a result of the infection. Additionally, no obesity-related markers, such as insulin, C-peptide, glucagon, leptin or cortisol were measured.

We demonstrated not only a reduction in gray matter volume associated with higher BMI in a cohort of post-COVID patients, but also link these changes, along with BMI, to distinct immunological changes, most pronounced in patients with cognitive deficits. This could help elucidate possible converging pathomechanisms of obesity and post-COVID condition in terms of chronic inflammation and structural brain changes. In the future, further research in this area could enable the development of specifically adapted immunomodulatory therapies for PCC in obese patients.

## Data Availability

All data produced in the present study are available upon reasonable request to the authors.

## Acknowledgement

We thank Ines Krumbein for overseeing MRI measurements and to Marie Troll, Antonia Toepffer, Lara Krickow, Eva-Maria Dommaschk, Maximilian Vollmer and Marlene Müller for MRI data collection and pre-analyzing blood samples.

